# Functional Magnetic Resonance Imaging of the amygdala and subregions at 3 Tesla: A scoping review protocol

**DOI:** 10.1101/2022.04.14.22273332

**Authors:** Sheryl L. Foster, Isabella A. Breukelaar, Kanchana Ekanayake, Sarah Lewis, Mayuresh S. Korgaonkar

## Abstract

**Background:** Functional Magnetic Resonance Imaging (fMRI) is a widely accepted and utilised method of investigating neural activation within the brain. There has been increasing awareness and understanding in the field of neuropsychology over the last 10-15 years that the amygdala plays an important role in many mental health conditions. Functional connectivity (FC) of the amygdala with other parts of the brain is well-documented in the literature; however the role of the amygdala and its reported connections is still not well understood and this can be attributed, in part, to its very small size. It is challenging to achieve adequate spatial resolution to visualise amygdala activation using 3T MRI systems that are in widespread use for this type of clinical research. Optimisation of protocols for improved data accuracy and reproducibility may potentially lead to standardisation and subsequent advancements in overall image quality in this field.

**Methods:** The protocol for this scoping review was developed in line with the Preferred Reporting Items for Systematic reviews and Meta-Analyses extension for Scoping Reviews (PRISMA-ScR) and registered with the Open Science Framework (OSF). A literature search of five databases (Medline, Embase, Web of Science, Google Scholar and Scopus) will be undertaken using a refined search strategy; peer-reviewed publications identified as being relevant will then be imported into Covidence software for abstract screening and data extraction by two reviewers working independently. The quantitative findings will be tabulated to provide an overview of the current methodologies for comparison. This will be accompanied by a narrative report summarising the extracted data in relation to the stated research questions.

**Discussion:** The objective of this scoping review is to identify and map the range of existing protocols used in fMRI for imaging the activation and FC patterns of the amygdala at 3 Tesla. This will be achieved by collating and presenting quantitative data relating to protocol parameter choices as well as other qualitative aspects of the data acquisition process.

**Registration:** Open Science Framework (OSF) – Registration type: OSF Pre-registration

Registration: https://osf.io/e3c28, DOI: 10.17605/OSF.IO/KW58P

## Background

Functional Magnetic Resonance Imaging (fMRI) is a widely accepted and utilised method of investigating neural activation within the brain (Glover, 2011). fMRI has advanced our understanding of the structure and function of distinct brain regions as well as how these different regions may be working together (Huettel, Song, & McCarthy, 2004). Although it has long been recognised by neuroscientists that the amygdala is an intrinsic component of the emotion circuitry of the brain (LeDoux, 2000), there has been increasing awareness and understanding in the field of neuropsychology over the last 10-15 years that the amygdala plays an important role in many mental health conditions (Leppänen, 2006; Schumann, Bauman, & Amaral, 2011).

The amygdalae comprises a pair of almond-shaped structures located deep in the temporal lobes of the brain with each amygdala being only around 10-20mm^3^ in size (Amunts et al., 2005). The brain has both structural and functional connections and investigation of the functional connectivity (FC) of the amygdala with other parts of the brain is well-documented in the literature; however the role of the amygdala and its reported connections is still not well understood (Mohanty, Sethares, Nair, & Prabhakaran, 2020) and this can be attributed, in part, to the very small size of the amygdala. Complicating matters further is that each amygdala is made up of three distinct subregions each with its own disparate connections to other parts of the brain (Bzdok, Laird, Zilles, Fox, & Eickhoff, 2013; Nieuwenhuys, Voogd, & van Huijzen, 2008).

Spatial resolution, the currency of MRI, is referenced on voxel volume and is a measure of pixel numbers in an imaging volume. The greater the number of pixels in a voxel, the higher the spatial resolution of an image; this is a requirement for very small structures such as the amygdala to be well-resolved. Although imaging with sufficiently high spatial resolution can be accomplished using high-field strength 7T systems that possess inherently higher signal-to-noise ratio (SNR), these systems are not widely available for clinical research (Iranpour, Morrot, Claise, Jean, & Bonny, 2015) and it is more challenging to achieve adequate spatial resolution to visualise amygdala activation using the lower strength 3T MRI systems that are in widespread use for this type of clinical research (Sladky et al., 2018).

There is reported evidence that higher resolution imaging protocols can distinguish additional activation patterns and connections in many brain areas including the amygdala, and this has been demonstrated at both 3T and 7T (Iranpour et al., 2015; Sladky et al., 2018). Additionally, some authors have reported imaging of amygdala activation using coronal and sagittal acquisition planes rather than the standard axial plane at 3T in order to improve SNR by reducing through-plane signal dephasing (Boubela et al., 2015; Kim, Somerville, Johnstone, Alexander, & Whalen, 2003). There is also evidence that different designs in radiofrequency (RF) coils used for signal reception can have significant implications for SNR and achievable spatial resolution (Gruber, Froeling, Leiner, & Klomp, 2018). Taking stock of the protocols in common use for fMRI data acquisition of the amygdala at 3T is the first step in potential optimisation and standardisation of protocols for improved data accuracy, reproducibility and overall quality.

### Study Rationale

Variability in imaging protocols, even at the same field strength, may result in significant disparity in data output and quality. In such a compact organ as the amygdala, this variability in protocols and data quality can have major implications for accurate reporting of activation and functional connections within the brain. To our knowledge, there is no apparent standardisation of fMRI protocols for imaging activation of the amygdala at 3T in the field of neuroimaging.

### Study Objective

The scoping review has been chosen as a method of summarising and disseminating findings in relation to 3T fMRI protocols in use for studying activation and functional connectivity (FC) in the amygdala and its subregions. The review will provide evidence as to current practice and document protocol disparities across clinical research facilities worldwide. It will potentially provide evidence and context for recommending that standardised and refined fMRI protocols are developed for reliably imaging activation of the amygdala. Optimised fMRI protocols applied consistently across imaging groups could lead to increased replication of findings and a greater understanding of the role of the amygdala in mental health and other conditions.

### Methodology

The protocol was developed in line with the Preferred Reporting Items for Systematic reviews and Meta-Analyses extension for Scoping Reviews (PRISMA-ScR)(Tricco et al., 2018) and registered with the Open Science Framework (OSF). The aim is to identify and chart specific fMRI protocol parameters and data acquisition techniques from a range of studies across multiple sites and geographic locations, the results of which will provide a transparent summary of current protocols for comparison and review by fMRI researchers. As such, this review will not extend to an appraisal of study quality.

The scoping review will follow the five steps proposed in the methodological framework of Arksey and O’Malley as outlined below (Arksey & O’Malley, 2005).

i. identifying the research question
ii. identifying relevant studies
iii. selection of eligible studies
iv. charting the data
v. collating, summarising and reporting the results

#### Step 1: Identifying the research question

The review question was developed in line with PCC (Population, Concept, Context) elements as outlined in the JBI Manual for Evidence Synthesis (Aromatis & Munn, 2020).

- Population – amygdala and subregions
- Concept – fMRI protocols
- Context – 3T

The main research question addressed in the scoping review is “What is the current 3 Tesla functional MRI protocol in research use for imaging activation of the amygdala and its subregions?”

There are five research sub-questions:

i. What was the value of the spatial resolution achieved (in mm^3^)
ii. What imaging plane was utilised?
iii. Was full brain coverage achieved?
iv. What was the sequence acquisition time?
v. What type of radiofrequency (RF) coil was utilised for signal reception?

#### Step 2: Identifying relevant studies

As per the recommendations of Peters and colleagues, the search strategy was developed using keywords based on the research question in partnership with co-authors (MK, SL, KE) (Peters et al., 2020). The latter co-author, an academic librarian, subsequently recommended appropriate and relevant databases in which to conduct the searches. An initial (more complex) search strategy was piloted together with the academic librarian, during which the electronic databases listed below were searched. The combination of databases selected was based on the findings of Bramer and colleagues who reported that the combination of the first four databases listed was optimal in systematic review literature searches with an overall recall rate of 98.3% (Bramer, Rethlefsen, Kleijnen, & Franco, 2017). Their recommendation of the addition of Scopus as a fifth database if the number of studies identified was low was also followed.

- Medline (M)
- Embase (E)
- Web of Science (WoS)
- Google Scholar (GS)
- Scopus (S)

The search strategy was then refined and simplified, following which it was retested on the Medline and Embase databases and results compared to those of the more complex strategy. As the simple search strategy (29M and 197E) identified more studies than the more complex strategy (16M and 118E), it was chosen for the review. Results of the electronic database searches will be presented in tabular format (Table 1).

**Table 1:**
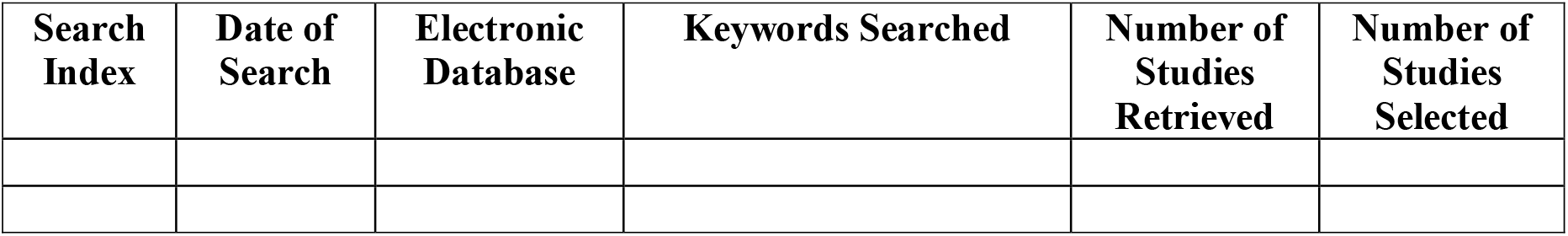
Electronic Database Search Recording Table.

Keywords or Medical Subject Headings (MeSH) terms for the search encompassed the following:

- Functional Magnetic Resonance Imaging OR functional MRI OR fMRI
- Amygdal* OR amygdal* nucleus
- Functional connect* OR FC (functional connectivity)
- 3 Tesla OR 3T

A hand search of the identified/selected references for inclusion will be performed to identify other potential studies that may have been missed. Any grey (unpublished) literature identified will be examined and its suitability assessed for inclusion.

#### Step 3: Selection of eligible studies

In accordance with the PCC framework outlined above, titles and abstracts will be screened for suitability. Further inclusion and exclusion criteria will also be applied in order to screen out irrelevant studies.

Inclusion Criteria

- Imaging performed at 3T only
- Studies including full fMRI parameter details for extraction in methodology section
- Clinical fMRI research protocols only – acceptable imaging times (exclude research studies seeking highest resolution possible at expense of translational value)

Exclusion criteria

- Animal studies
- Qualitative studies, reviews, and conference abstracts
- Full-text studies that could not be sourced for review

Valuable information regarding paediatric protocols may be identified for comparison in the review; therefore no age limits have been specified. Similarly, a decision was made not to filter out studies in other languages at the screening stage in the interests of inclusivity; it is important to examine current practices as globally as possible. It was also deemed unnecessary to exclude studies based on a year of publication as 3 Tesla MRI systems only became clinically available in the early 2000s.

The search strategy has been refined and the full database searches will be undertaken. Identified articles will be imported into Covidence software (Version 2745 34609193)(Covidence, 2022). Duplicates will be identified and cross checked prior to removal in readiness for the screening process. The title and abstract screening process will be conducted by two authors (SF and IB) during April and May, 2022, following which the same authors will perform full-text screening of the studies selected for inclusion. An academic librarian will assist in obtaining full-text articles not freely available and any unresolved differences of opinion between the two reviewers as to study eligibility will be resolved by a third reviewer and reported in the review.

The recommendations outlined in the Preferred Reporting Items for Systematic Reviews and Meta-Analyses Extension for Scoping Reviews (PRISMA-ScR) checklist (Tricco et al., 2018) will be followed and the search and screening strategy will be charted for reporting the results for publication (Figure 1).

**Figure 1:**
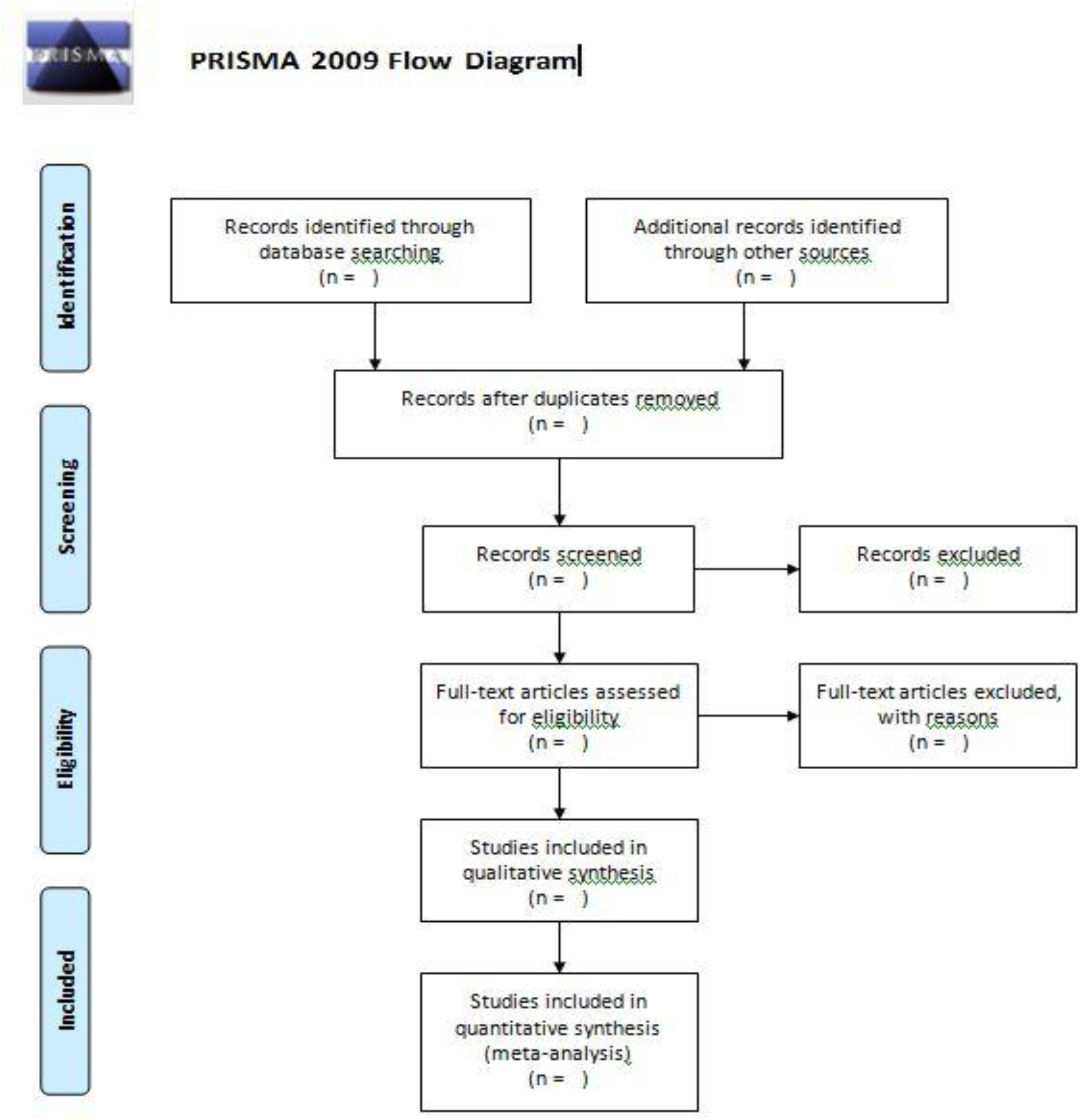
PRISMA 2009 flow diagram (Moher, Liberati, Tetzlaff, Altman, & The, 2009)

#### Step 4: Charting the Data

In order to determine the variables for extraction a framework for charting and assessment of the data has been developed in line with the recommendations of Tricco and colleagues (Tricco et al., 2018). Prior to full data extraction, two reviewers will test the suitability of the charting framework on 10% of the included studies and discuss any revisions required prior to implementation. Following this process, the two reviewers, working independently to reduce the risk of bias, will use the final version of the data charting form to capture electronically the fields listed in Table 2.

**Table 2:** Data extraction framework.

| Main Category | Description |
| --- | --- |
| Authors |  |
| Title |  |
| Journal |  |
| Publication Year |  |
| Type of Publication | specify whether website, journal, original research |
| Study Setting | specify the geographical area/healthcare or imaging setting |
| Study Population & Sample Size | specify age and health condition under investigation |
| Methodology (quantitative) | specify fMRI imaging parameters relating to spatial resolution (matrix size and slice thickness) and sequence acquisition time in minutes and seconds |
| Methodology (qualitative) | specify imaging plane, RF coil selection, brain coverage, sequence type |
| Offline Data Analysis Method | specify software programs used |
| Reported Outcomes | describe relevant study findings and any limitations |

#### Step 5: Collating, summarising and reporting the results

The quantitative findings will be tabulated as a means of reporting an overview of the current methodologies for comparison. This will be accompanied by a narrative report summarising the extracted data in relation to the following outcomes; spatial resolution of the fMRI protocol in mm^3^, plane of imaging, the type of RF coil employed and whether whole brain coverage was achieved. These results, reported together, will provide an accessible record of current practices in relation to the research question for the benefit of specialised fMRI researchers.

## Discussion

The objective of this scoping review is to identify and map the range of existing protocols used in fMRI for imaging the activation and FC patterns of the amygdala at 3 Tesla; this will be achieved by collating and presenting quantitative data relating to protocol parameter choices as well as other qualitative aspects of the data acquisition process.

A preliminary limited review of the relevant literature has revealed a range of parameter selections in use in research protocols investigating the functional connectivity of the amygdala resulting in widely disparate spatial resolution values. To our knowledge this review will be the first to highlight these protocol inconsistencies in a structured and overarching manner and it likely has the potential to generate discussion and bring about review and advancement of practice in this highly specialised field.

A limitation of this review is that, due to its scoping nature, the quality of the data produced by the protocols identified will not be assessed; however the quantitative nature of the reported data together with the collated information regarding the qualitative data may prove to be a sound basis for discussion and further research built on the findings of this scoping review.

## Data Availability

All data produced in the present work are contained in the manuscript

## Abbreviations

fMRI: functional Magnetic Resonance Imaging;
3T: 3 Tesla
RF coil: radiofrequency coil
PRISMA-ScR: Preferred Reporting Items for Systematic Reviews and Meta-Analyses Extension for Scoping Reviews
OSF: Open Science Framework
PCC: Population, Concept, Context
MeSH: Medical Subject Headings
FC: Functional Connectivity

## Authors’ contributions

SF conceptualized the study and prepared the draft protocol under the supervision of MK and SL. SF, MK, SL and IB contributed to the development of the background, design of the study, and planned output of the research. KE advised on database selection and searches. SF and IB prepared the manuscript, and all authors reviewed, edited and approved the final manuscript for submission.

## Availability of data and materials

All data generated or analysed during this study will be included in the published scoping review manuscript

## Ethics approval and consent to participate

Not applicable

## Consent for publication

Not applicable

## References

Amunts, K., Kedo, O., Kindler, M., Pieperhoff, P., Mohlberg, H., Shah, N. J., … Zilles, K. (2005). Cytoarchitectonic mapping of the human amygdala, hippocampal region and entorhinal cortex: intersubject variability and probability maps. Anatomy and Embryology, 210(5), 343–352. doi:10.1007/s00429-005-0025-5

Arksey, H., & O’Malley, L. (2005). Scoping studies: towards a methodological framework. International Journal of Social Research Methodology, 8(1), 19–32. doi:10.1080/1364557032000119616

Boubela, R. N., Kalcher, K., Huf, W., Seidel, E.-M., Derntl, B., Pezawas, L., … Moser, E. (2015). fMRI measurements of amygdala activation are confounded by stimulus correlated signal fluctuation in nearby veins draining distant brain regions. Scientific Reports, 5(1), 10499. doi:10.1038/srep10499

Bramer, W. M., Rethlefsen, M. L., Kleijnen, J., & Franco, O. H. (2017). Optimal database combinations for literature searches in systematic reviews: a prospective exploratory study. Systematic reviews, 6(1), 245. doi:10.1186/s13643-017-0644-y

Bzdok, D., Laird, A. R., Zilles, K., Fox, P. T., & Eickhoff, S. B. (2013). An investigation of the structural, connectional, and functional subspecialization in the human amygdala. Human Brain Mapping, 34(12), 3247–3266.

Covidence. (2022). Covidence systematic review software. from Veritas Health Innovation http://www.covidence.org

Glover, G. H. (2011). Overview of functional magnetic resonance imaging. Neurosurgery clinics of North America, 22(2), 133–vii. doi:10.1016/j.nec.2010.11.001

Gruber, B., Froeling, M., Leiner, T., & Klomp, D. W. J. (2018). RF coils: A practical guide for nonphysicists. Journal of magnetic resonance imaging : JMRI, 48(3), 590–604. doi:10.1002/jmri.26187

Huettel, S. A., Song, A. W., & McCarthy, G. (2004). Functional magnetic resonance imaging (1st ed. ed.). Sunderland, Mass: Sinauer Associates.

Iranpour, J., Morrot, G., Claise, B., Jean, B., & Bonny, J.-M. (2015). Using high spatial resolution to improve BOLD fMRI detection at 3T. PLoS ONE, 10(11), e0141358. doi:10.1371/journal.pone.0141358

Kim, H., Somerville, L. H., Johnstone, T., Alexander, A. L., & Whalen, P. J. (2003). Inverse amygdala and medial prefrontal cortex responses to surprised faces. Neuroreport, 14(18), 2317–2322. doi:10.1097/00001756-200312190-00006

LeDoux, J. E. (2000). Emotion Circuits in the Brain. Annual Review of Neuroscience, 23(1), 155–184. doi:10.1146/annurev.neuro.23.1.155

Leppänen, J. M. (2006). Emotional information processing in mood disorders: a review of behavioral and neuroimaging findings. Current Opinion in Psychiatry, 19(1).

Mohanty, R., Sethares, W. A., Nair, V. A., & Prabhakaran, V. (2020). Rethinking Measures of Functional Connectivity via Feature Extraction. Scientific Reports, 10(1), 1298. doi:10.1038/s41598-020-57915-w

Moher, D., Liberati, A., Tetzlaff, J., Altman, D. G., & The, P. G. (2009). Preferred Reporting Items for Systematic Reviews and Meta-Analyses: The PRISMA Statement. PLOS Medicine, 6(7), e1000097. doi:10.1371/journal.pmed.1000097

Nieuwenhuys, R., Voogd, J., & van Huijzen, C. (2008). Telencephalon: amygdala and claustrum. The human central nervous system, 401–426.

Peters, M. D. J., Marnie, C., Tricco, A. C., Pollock, D., Munn, Z., Alexander, L., … Khalil, H. (2020). Updated methodological guidance for the conduct of scoping reviews. JBI Evid Synth, 18(10), 2119–2126. doi:10.11124/jbies-20-00167

Schumann, C. M., Bauman, M. D., & Amaral, D. G. (2011). Abnormal structure or function of the amygdala is a common component of neurodevelopmental disorders. Neuropsychologia, 49(4), 745–759. doi:10.1016/j.neuropsychologia.2010.09.028

Sladky, R., Geissberger, N., Pfabigan, D. M., Kraus, C., Tik, M., Woletz, M., … Windischberger, C. (2018). Unsmoothed functional MRI of the human amygdala and bed nucleus of the stria terminalis during processing of emotional faces. NeuroImage, 168, 383–391.

Tricco, A. C., Lillie, E., Zarin, W., O’Brien, K. K., Colquhoun, H., Levac, D., … Straus, S. E. (2018). PRISMA Extension for Scoping Reviews (PRISMA-ScR): Checklist and Explanation. Annals of Internal Medicine, 169(7), 467–473. doi:10.7326/M18-0850

